# *CFTR* Gene Variant Detection in Moroccan Individuals via Nanopore Long-Read Sequencing

**DOI:** 10.1101/2025.02.14.25322057

**Authors:** Nada El Makhzen, Alexander Nater, Jean-Sébastien Rougier, Alexandre Bokhobza, Javier Sanz, Christiane Zweier, Anne-Flore Hämmerli, Rémy Bruggmann, Laila Bouguenouch, Mounia Lakhdar Idrissi, Hugues Abriel

## Abstract

**Background:** Cystic fibrosis (CF) is an autosomal recessive disease resulting from pathogenic CF transmembrane conductance regulator (*CFTR*) pathogenic gene variants. While CF’s frequency varies among ethnicities, its epidemiology, clinical manifestations, and mutational profiles in Africa still must be explored due to the absence of a comprehensive public health strategy there. This study postulates that complete sequencing of *CFTR* using Oxford Nanopore Technology (ONT)-based long-read sequencing enhances the diagnostic yield.

**Methods:** To amplify ∼25-kb fragments covering the whole *CFTR* gene (NM_000492.4), we designed 11 primer pairs, and barcoded libraries were prepared and sequenced on ONT flow cells (R10.4.1) using an Mk1C device. Variant pathogenicity was assessed by expressing the variant channel in HEK293 cells and examining expression through immuno-blotting.

**Results:** With sequencing data obtained from 9 Moroccan individuals (6 probands with suspected CF diagnoses and 3 parents), we identified the following variants: c.680T>G p.Leu227Arg, c.1521_1523del p.Phe508del, c.3484C>T p.Arg1162*, c.1090T>C p.Ser364Pro, c.3233T>C p.Phe1078Ser and c.2991G>C p.Leu997Phe. The analytical pipeline we developed allowed the phasing of the variants. Sanger sequencing confirmed all these results. The previously uncharacterised *CFTR* variants p.Ser364Pro and p.Phe1078Ser exhibit diminished expression in HEK293 cells, substantiating their pathogenic nature, with p.Phe1078Ser responding positively to the *in vitro* treatment with CFTR-modulator molecules.

**Conclusions:** This study demonstrates the potential of long-read sequencing using ONT as an efficient means to detect CF-causing variants in African populations. Given the significant genetic heterogeneity in Africa, this technique can serve as an affordable molecular screening tool for CF, especially in areas with constrained access to genetic screening.

**Highlights:**

- Here, we demonstrate the efficiency of sequencing the complete *CFTR* gene using Oxford Nanopore Technology, an information-rich and accurate method.
- Not-yet-reported *CFTR* variants of unknown significance were identified from a small cohort of nine cystic fibrosis patients and parents from Fez, Morocco.
- The biochemical characterisation of the *CFTR* variants p.Ser364Pro, p.Phe1078Ser, and p.Leu997Phe highlights the importance of conducting expression studies when genetic variants are identified.
- Our proposed approach will enhance diagnostic outcomes, particularly in populations with significant genetic diversity, such as those in Africa.

## 1. Introduction

Cystic fibrosis (CF) is an autosomal recessive disease caused by pathogenic variants in the ∼250-kb CF transmembrane conductance regulator (*CFTR*) gene, which affects the function of the ion channel protein to maintain chloride balance across epithelial cells. More than 2000 variants have been identified in the *CFTR* gene, with more than 700 causing CF (HGMD Professional 2023.4, CFTR2 (www.cftr2.org)), which is characterised by chronic lung disease, pancreatic insufficiency, elevated sweat chloride concentration levels, and obstructive azoospermia [1]. CF is a significant global health concern that affects individuals worldwide; however, many African patients continue to experience misdiagnosed CF, preventing them from receiving adequate medical care. This is mainly because many African hospitals rarely perform screening sweat tests. In addition, molecular genetic tests necessary to diagnose CF are costly and mostly unavailable in many African countries, leading to a lack of accurate and timely CF diagnosis. Implementing sequencing as the primary diagnostic strategy may be appropriate for many low- and middle-income countries [2]. The time and costs associated with diagnosing CF may significantly decrease by integrating modern DNA sequencing technologies with suitable bioinformatics pipelines [3].

Developing next-generation sequencing (NGS) technologies has improved biomedical research and significantly increased the output of sequencing data. However, several studies have shown limitations that may impact its accuracy for diagnostic purposes [4], [5]; the short-read length (∼150 bp) is the most noticeable constraint for NGS [5]. Although short-read sequencing dominated NGS until recently, there are new sequencing platforms that can produce long multi-kilobase reads. These later platforms allow for long-range haplotype creation and reference-free genomic assembly [6]. Long-read sequencing (LRS) technology may accurately characterise genetic variation and regions that are challenging to evaluate with current short-read NGS. Nevertheless, compared with short-read NGS, LRS also has limitations and challenges related to the quality of the initial material, error rates, and costs [7]. For example, DNA extraction methods and high-molecular-weight DNA handling still need improvement. In addition, raw data processing, mapping, and variant calling tools are less developed for LRS than short-read NGS [7]. Consequently, LRS has mainly been applied at this stage to look into genetic diseases with known or highly suspected disease loci [7].

In this study, we evaluate the feasibility and efficiency of using LRS from Oxford Nanopore Technology (ONT) for complete sequencing of the ∼250-kb *CFTR* gene, encompassing both coding and non-coding regions and to identify variants responsible for CF in African populations, initially with a focus on nine individuals from Morocco. LRS with ONT effectively identified CF-causing variants. Furthermore, we present evidence for the likely pathogenic nature of the previously uncharacterised *CFTR* variants p.Ser364Pro and p.Phe1078Ser found in two Moroccan CF patients.

## 2. Materials and Methods

### 2.1. Study participant recruitment and sample processing

Fifteen individuals were referred by pediatricians from the University Hospital Hassan II (Sidi Mohamed Ben Abdellah University, Fez, Morocco) to its Laboratory of Medical Genetics and Oncogenetics. Of them, 9 were selected, including 6 probands with suspected CF and 3 healthy parents. Individual data were collected from the patient’s medical files, including each patient’s age at the onset of symptoms and upon diagnosis, as well as place of origin and presence of consanguinity. If available, we documented the *CFTR* variants and sweat test results. In addition, we assessed the results of various clinical examinations, including imaging, microbiology, and lung function tests (if they had been conducted), as well as the respiratory diseases that occurred throughout surveillance. Blood samples were then collected from those who consented to participate in the study. These samples were sent to the Medical Genetics Lab for DNA extraction and forwarded to the Ion Channels and Channelopathies Laboratory at the Institute for Biochemistry and Molecular Medicine (University of Bern, Switzerland) for LRS using ONT (Figure 1). The Committee of Ethics Biomedical Research, Mohammed V University, Rabat, Morocco approved the current study.

**Figure 1:**
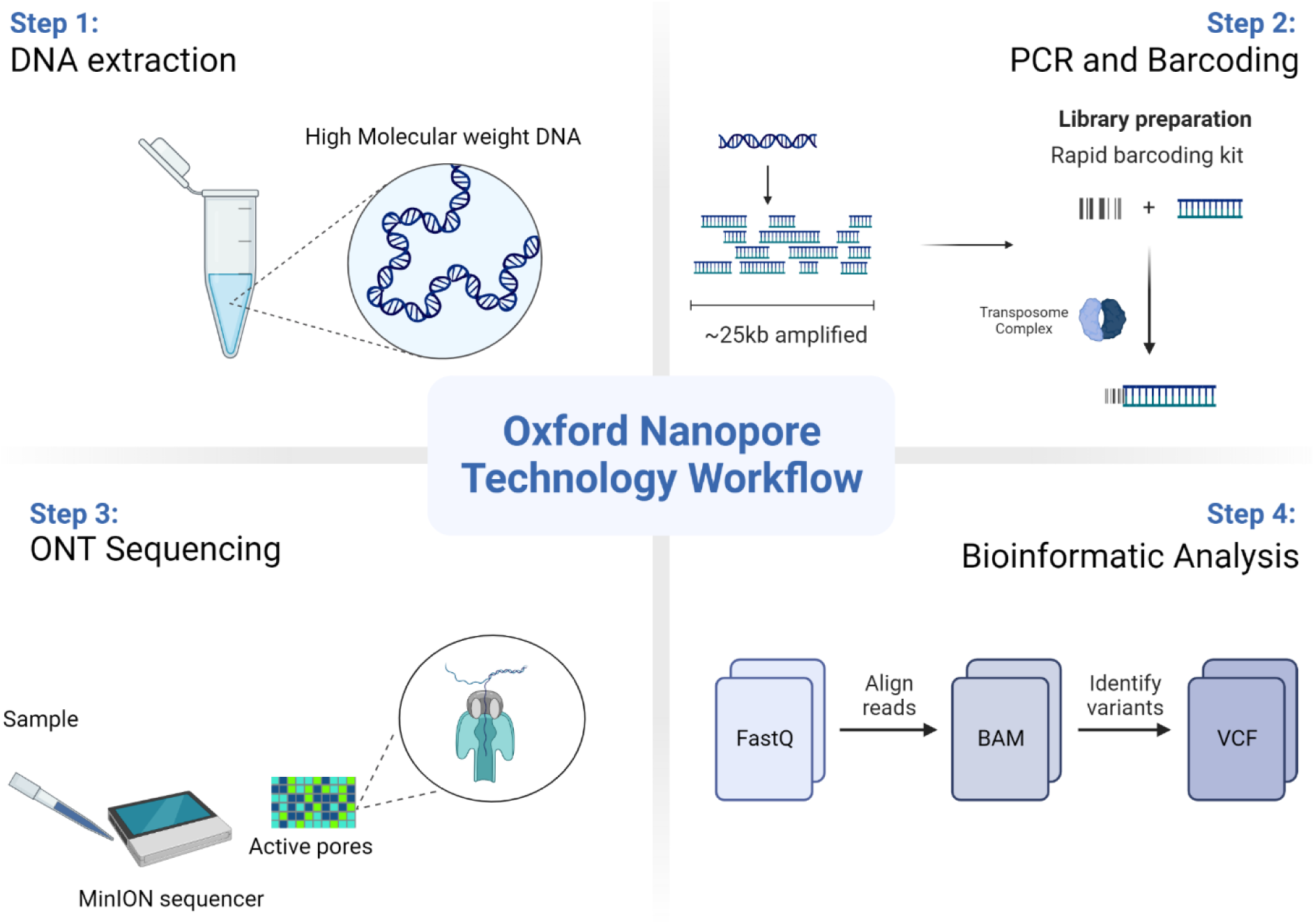
Experimental overview. Visualisation of the Oxford Nanopore Technology (ONT) sequencing workflow using the MinION device.

### 2.2. Generation of sequence data

- A detailed protocol has been published on protocols.io <u>(**10.17504/protocols.io.ewov1o7y7lr2/v1**)</u>. (Private link for reviewers: https://www.protocols.io/private/BA87210BA79D11EDB1C50A58A9FEAC02 to be removed before publication.)

### 2.3. Data Analysis and Interpretation

Unless otherwise stated, default parameters were used for all bioinformatics tools. We converted fast5 output from the sequencing to pod5 format and performed offline base-calling with Dorado v0.8.3 using the ‘sup’ model [8]. We performed quality control on the resulting FASTQ files using FastQC v0.12.1 and NanoPlot v1.41.6 [9]. The ONT reads were filtered with fastp v0.23.4 to remove low-quality reads (<40% of bases with base quality <15) [10] and aligned to the GRCh38 human reference genome using minimap2 v2.28 with the ‘lr:hq’ preset optimised for Q20+ ONT reads [11]. We calculated mapping and depth statistics with the SAMtools v1.20 *flagstat* command and Mosdepth v0.3.8 [12], [13]. Variant calling for single-nucleotide and insertion/deletion variants (SNVs and indels) was performed for each sample with Clair3 v1.0.10 [14] using BAM files created in the mapping step. The genomic VCF (gVCF) files generated in the variant calling were then used to jointly call genotypes across the entire cohort using GLnexus v1.4.1 with the configuration file provided by Clair3 [15].

We used a two-step approach to phase the joint VCF file since the length of the overlaps between amplicons did not allow for read-based phasing across the entire *CFTR* gene. In the first step, we applied reference-based phasing with Eagle2 v2.4.1 [15] using a panel from the 1000 Genomes Project of 3,202 individuals sequenced to 30x depth [16]. This allowed us to phase common variants found in both the reference panel and our cohort, resulting in a phasing backbone across the *CFTR* gene. To phase the rare variants not included in the reference panel, we applied a read-based phasing approach with WhatsHap v2.3 using the phased VCF output from the previous step together with the mapped ONT reads [17]. This resulted in phasing blocks spanning the entire *CFTR* gene for all 9 individuals.

We called large structural variants (SV) from the mapped ONT reads using Sniffels2 v2.4 [18] and cuteSV v2.1.1 [19]. For Sniffles2, we first called SV candidates in each individual separately, followed by a population-level calling step across the entire cohort using the SNF files from the first step as input. For cuteSV, we called each individual separately using the recommended settings for ONT data, followed by a merging of sample-wise VCF files with SURVIVOR v1.0.7 [20] (1 kb maximum breakpoint distance, minimum support of 1, accounting for SV type and strand). We then force-called the SVs in the merged VCF file in each individual using cuteSV in genotype mode and merged the resulting sample-wise VCF files again into a population-level VCF file with SURVIVOR.

Ensembl Variant Effect Predictor (VEP) [21] was used to annotate the phased VCF files. Aligned reads were visualised using the Integrative Genomics Viewer (IGV) [22]. Variants with an allele frequency >1% in public databases (gnomAD V4.0, dbSNP) were removed from further analysis. The remaining variants were prioritised based on the presence in variant databases (Clinvar, HGMD Professional 2023.4, CFTR2 (www.cftr2.org)), allele transmission (e.g., biallelic variants), and predicted variant severity. Candidate variants were predicted to be deleterious by multiple in silico prediction tools (Polyphen-2, SIFT). The variant classification was performed according to CFTR2 or the criteria of the American College of Medical Genetics and Genomics (ACMG) [23].

### 2.4. Sanger Sequencing for the validation experiments

*CFTR* variants detected by ONT were confirmed by Sanger sequencing. The corresponding *CFTR* exons were amplified using genomic DNA as a template. They were sequenced on an ABI Prism 3500XL Genetic Analyzer (Applied Biosystems, Foster City, CA, USA) using BigDye Terminator v3.1 chemistry (Applied Biosystems, Foster City, CA, USA). Sequence data were analysed with Sequence Pilot v4.3 (JSI Medical Systems, Ettenheim, Germany).

### 2.5. Biochemical characterisation of the CFTR variants

Human embryonic kidney 293 (HEK-293) cells were cultured at 37°C with 5% CO_2_ in DMEM medium (Gibco, Fisher Scientific, Zurich, Switzerland), supplemented with 4 mM Glutamine, 10% FBS and a cocktail of streptomycin–penicillin antibiotics. HEK-293 cells were transiently transfected with 5 µg cDNA plasmids of interest in 100-mm petri dishes using LipoD293 (TebuBio company) in Vitro DNA Transfection Reagent. The cDNA plasmids of interest were p.WT-CFTR (wild-type), p.Phe508del-CFTR (a generous gift from Prof. Becq, Poitiers, France), p.Ser364Pro-CFTR, p.Leu997Phe-CFTR, and p.Phe1078Ser-CFTR (outsourced to GenScript company). Following incubation of 48 h at 37 °C with 5 % CO_2_, cells expressing CFTR variants were washed twice with cold 1X PBS and were incubated with 4 ml of 0.5 mg/ mL of EZlinkTM Sulfo-NHS-SS-Biotin (Thermo Scientific, Waltham, MA, USA) in cold 1X PBS for 30 min at 4 °C. Subsequently, the cells were washed twice with 200 mM Glycine in cold 1X PBS and once with cold 1X PBS to inactivate and remove the excess biotin, respectively. The cells were then lysed for 1 h at 4°C with 1X lysis buffer (50 mM HEPES, 150 mM NaCl, 1.5 mM MgCl_2_, 1 mM EGTA (pH 8); 10% glycerol, 10% Triton X-100, 1X Complete Protease Inhibitor Cocktail). Cell lysates were centrifuged at 16,100 rcf at 4°C for 15 min. A Bradford assay method on 590 nm was then performed using the Glomax (Promega) Plat Reader System (Switzerland) to determine the protein concentration of the supernatant. One point five mg of total protein was incubated with 50 μL Streptavidin Sepharose High-Performance beads (GE Healthcare, Uppsala, Sweden) for 2 h at 4 °C, and the remaining supernatant was kept as the input. The beads were washed four times with 1X lysis buffer before elution and incubation with 50 μL of LDS sample buffer (Invitrogen) and 100 mM DTT for 30 min at 37 °C. These biotinylated fractions were analysed as CFTR expressed at the cell surface. The input fractions, analysed as the total expression of CFTR, were resuspended with LDS Sample Buffer plus 100 mM DTT to give a concentration of 1 mg/ mL and incubated at 37 °C for 30 min.

Protein samples were loaded on 8% Tris polyacrylamide gradient gels, transferred with the Trans-Blot Turbo Transfer System (Bio-Rad, Cressier, Switzerland), blocked for 1 h at room temperature with 5 % milk in 1X TBS and detected with the following antibodies: 1) primary rabbit anti-hCFTR (R&D systems, cat #MAB25031), anti-α-actin (Sigma, cat #A2066) and mouse anti-α-1 Na^+^/K^+^ ATPase (Abcam ab7671), as a loading control, in a 1:1000 dilution in 1 % BSA and TBS+ 0.1 % Tween, and 0.02% Sodium-azide) secondary antibodies IR Dye 800 CW, anti-rabbit diluted (1:20,000) in TBS + 0.1% Tween and IR Dye 700 CW, anti-mouse diluted (1:20,000). All scans were taken using the Fusion FX7 device (Witec AG, Sursee, Switzerland). Total CFTR protein expression is composed of band C, which corresponds to the mature fully-glycosylated form of CFTR (170 kDa), and band B, which corresponds to the core-glycosylated form (150 kDa); CFTR was quantified using the Fusion FX7 (Witec AG) tool and normalised to the loading control protein, α-1 Na+/K+ pump ATPase. Three independent experiments were performed. GraphPad Prism Software (version 9.5.1, La Jolla, CA, USA) was used for statistical analysis. The unpaired ANOVA was used to determine the statistical significance of the results, presented as mean ± SEM with *p*<0.05.

### 2.6. Biotinylation assay – CFTR modulator drug treatment of the CFTR variants p.Ser364Pro, and p.Phe1078Ser

To characterize CFTR variants of uncertain significance, p.Ser364Pro and p.Phe1078Ser, for their responsiveness to the CFTR modulator molecules—two correctors (elexacaftor and tezacaftor) and the potentiator ivacaftor—HEK293 cells were cultured and transfected following the manufacturer’s instructions described in the previous step, with the cDNA plasmids of interest. Each transfection was done twice to have two groups: a treated group with CFTR modulator drugs and a control group treated with DMSO. Six hours post-transfection, the transfected cells were treated with the drugs at the following drug concentration: elexacaftor: 8.5 µM, tezacaftor: 5 µM, and ivacaftor: 10 µM for 12 hours. Biotinylation of the cells was conducted 24 h post-treatment following the manufacturer’s instructions previously described.

## 3. Results

### 3.1. African-Moroccan patients’ sequencing results

We analysed the coding and noncoding regions of the complete *CFTR* gene from 9 Moroccan-African individuals (6 probands with suspected CF diagnoses and 3 parents) using ONT amplicon sequencing (Figure 3).

Three of the probands had positive sweat test results (chloride concentration>110 mmol/L), one had an intermediate value, one was negative, and one had no sweat test (see Table 1). Specifically, using the variant-filtering approach described in the methods sections, we identified three CF-causing variants, two variants of uncertain significance (VUS), and one non-CF-causing variant in the heterozygous state (Table 1). Notably, during genetic analysis, a CF patient (F1-II1) was identified as having two CF-causing variants, p.Phe508del and p.Leu227Arg. This patient later had a positive sweat test confirming this result, with a chloride concentration of 117 mmol/L. Furthermore, screening of the parents (F1-I1 and F1-I2) confirmed the presence of these variants and thus compound heterozygosity. The phasing efficiency was confirmed using this trio and was proved to be consistent over the majority of the *CFTR* gene, except for two apparent switch errors in (F1-I2) towards the end of the gene area (7:117,626,788-117,627,457 and 7:117,671,063-117,671,069). Additionally, a VUS, p.Ser364Pro (Figure 2), was identified in combination with the CF-causing variant p.Arg1162* in a patient (F2-II1) who was hospitalised with respiratory issues and exocrine pancreatic insufficiency. This patient also had a positive sweat test with a chloride concentration of 113 mmol/L. Along with the CF-causing variant p.Arg1162*, we identified another VUS, p.Phe1078Ser, in a patient (F6-II1) who was hospitalised at a young age with recurrent respiratory infections with a positive sweat test and a chloride concentration of 111 mmol/L. Due to a lack of parental samples, compound heterozygosity could not be confirmed in the latter two individuals. One proband (F3-II1) diagnosed with mild CF symptoms, along with a negative sweat test result, was identified as having one known CF-causing variant, p.Arg1162*, see Table 1, in a heterozygous state. This observation raises the possibility that this proband is a *CFTR*-related disorder patient. For the other 2 individuals (F5-I2, and F5-II1) no pathogenic variants were detected with our sequencing. All the sequencing results obtained using the new ONT protocol were confirmed using traditional Sanger sequencing at the accredited genetics laboratory at the University Hospital of Bern.

**Figure 2:**
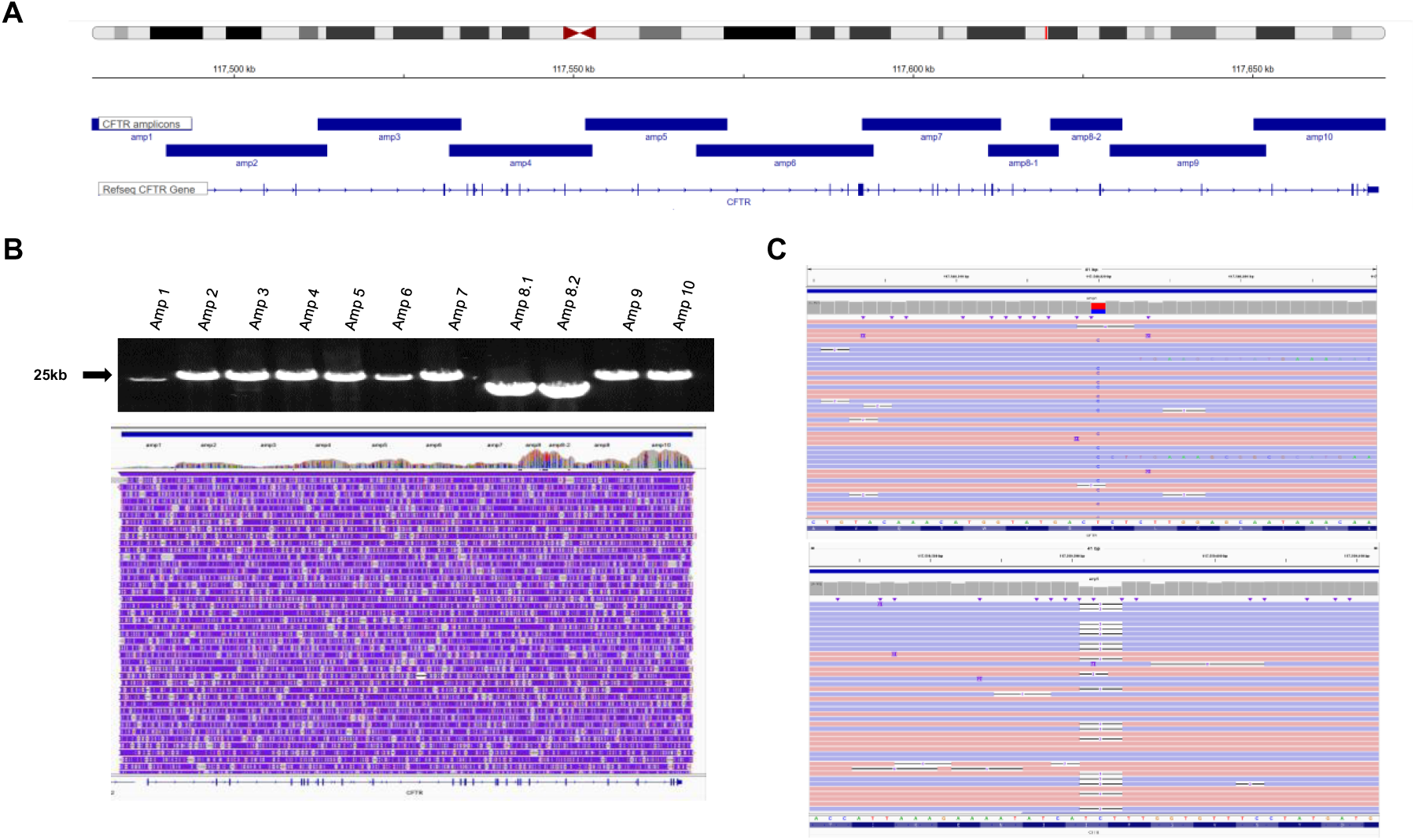
(A) Protocol illustration for amplifying the ∼250-kb *CFTR* gene using 11 long-range PCRs, with amp8 being split into two amplicons. (B) This panel demonstrates the amplification of the 11 amplicons and their subsequent sequence alignment via ONT long-read technology. (C) IGV visualisation of the variant *CFTR* c.1090T>C p.Ser364Pro, and *CFTR* c.1521_1523del p.Phe508del in the heterozygous state (https://igv.org/app/).

**Figure 3:**
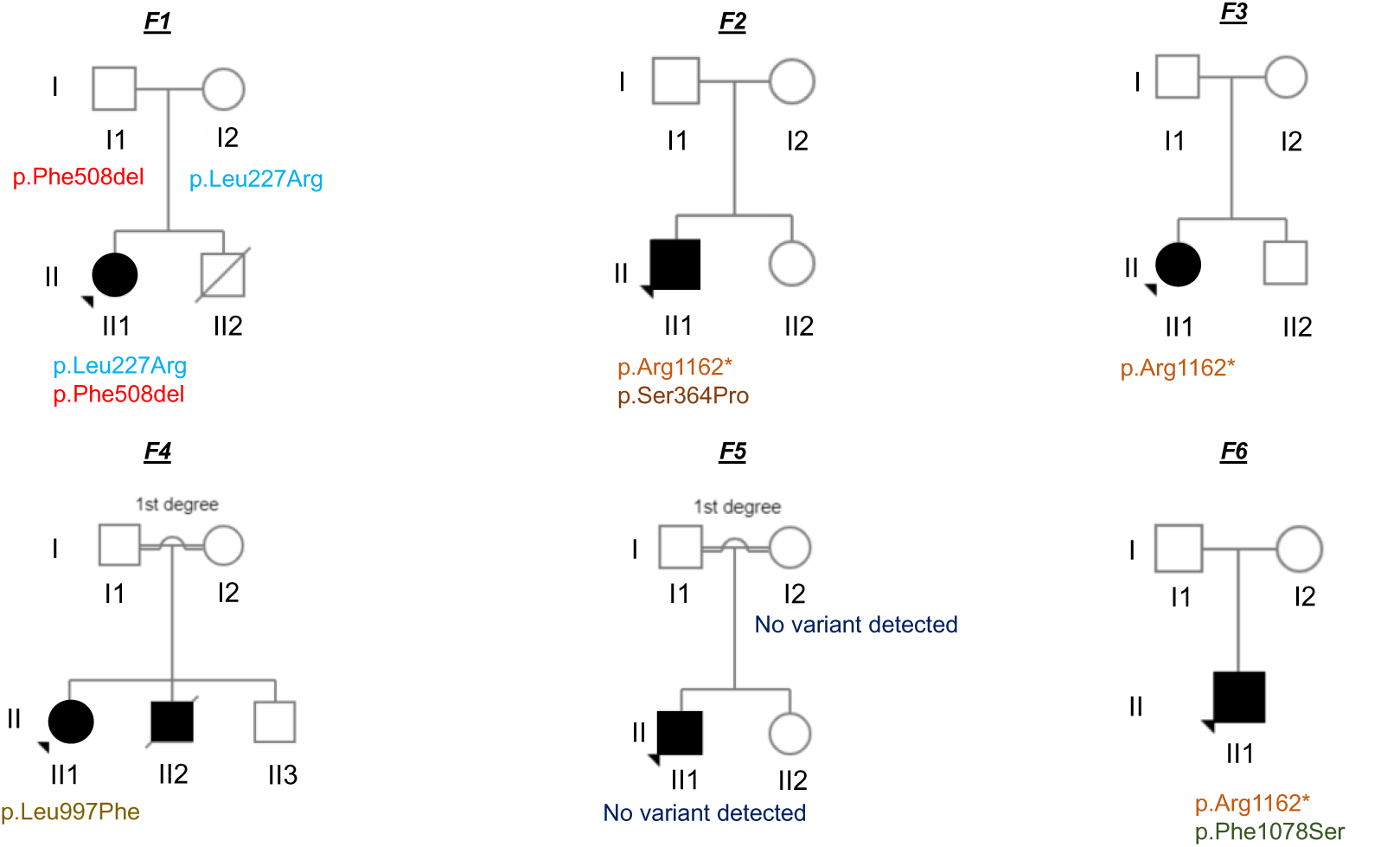
Pedigrees representing the relationship of the 9 sequenced individuals (6 probands with suspected CF and 3 parents).

**Table 1:**
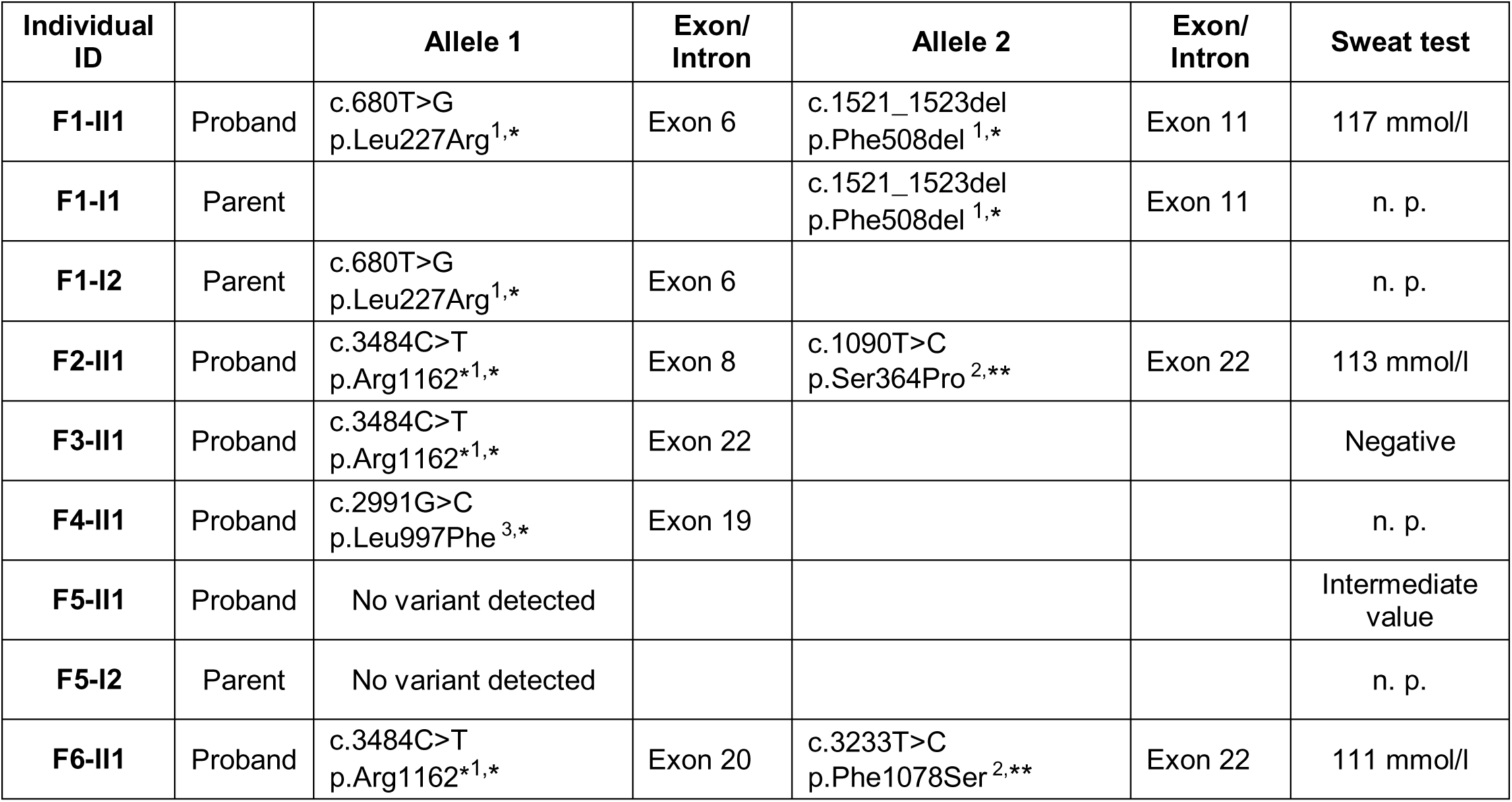
Detected genetic variants, n. p. = not performed, * Classification according to CFTR2 (www.cftr2.org), ** Classification according to ACMG guidelines. ^1^CF-causing, ^2^variant of uncertain significance, ^3^Non-CF causing.

### 3.2. Biochemical characterisation of the *CFTR* variants p.Ser364Pro, p.Leu997Phe, and p.Phe1078Ser

To assess their possible pathogenicity, we analysed the total cellular expression of *CFTR* variants p.Ser364Pro, p.Leu997Phe, and p.Phe1078Ser, which had not been characterised previously (Table 1) after transient transfection of HEK293 cells and its expression at the cell surface by immuno-blotting and cell surface biotinylation experiments. The immuno-blotting results (Figure 4) indicate a significant decrease in whole-cell expression of the p.Ser364Pro and p.Phe1078Ser CFTR variants when compared with WT, except for the p.Leu997Phe variant, which has a similar expression as the WT. The reduced expression was similar to that observed with the positive control, the p.Phe508del variant, which is known to be significantly less expressed than its WT counterpart. These findings suggest that the p.Ser364Pro and p.Phe1078Ser variants identified in Moroccan CF patients are likely pathogenic for CF rather than of unknown significance.

**Figure 4:**
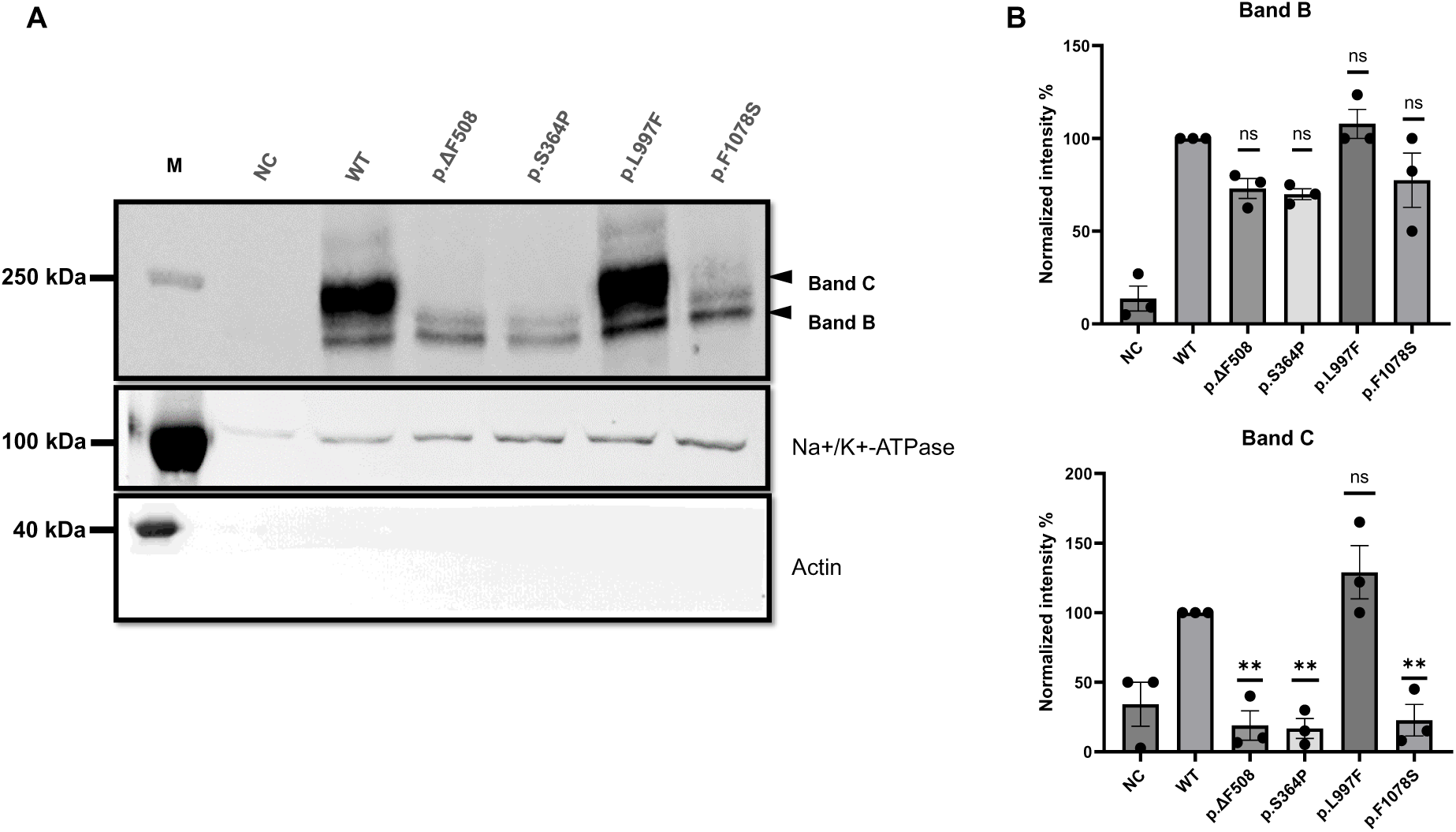
**(A)** Immuno-blots of biotinylated experiments showing the expression of p.CFTR-WT (WT) and the following 4 variants: p.Phe508del (p.ΔF508), p.Ser364Pro (p.S364P), p.Leu997Phe(p.L997F), and p.Phe1078Ser (p.F1078S). The variants p.Ser364Pro, and p.Phe1078Ser show a loss of expression that is at least as important compared with p.Phe508del. **(B)** Quantification of three different blots performed as (A). Data are presented as mean ± SEM; \**p*<0.05; \*\**p*<0.01; \*\*\**p*<0.001; \*\*\*\**p*<0.0001; ns: non-significance, with p-value representing WT VS variant. M: Marker, NC: negative control.

### 3.3. Biotinylation assay – CFTR modulator drug treatment of the CFTR variants p.Ser364Pro, and p.Phe1078Ser

We then assessed the *in vitro* effects of two correctors (elexacaftor and tezacaftor) and the potentiator drug ivacaftor. After 12-hour concomitant treatment with these three molecules, we analysed the surface expression of the CFTR variants p.Ser364Pro and p.Phe1078Ser by isolating biotinylated proteins and performing immuno-blot analysis. The results (Figure 5) showed a significant increase in the surface expression of the p.Phe1078Ser variant, as indicated by an increase in the intensity of the mature band C in the biotinylated fraction compared to the WT and p.Phe508del, known to be positively regulated by these compounds (Figure 5). In contrast, the p.Ser364Pro is insensitive to this treatment.

**Figure 5:**
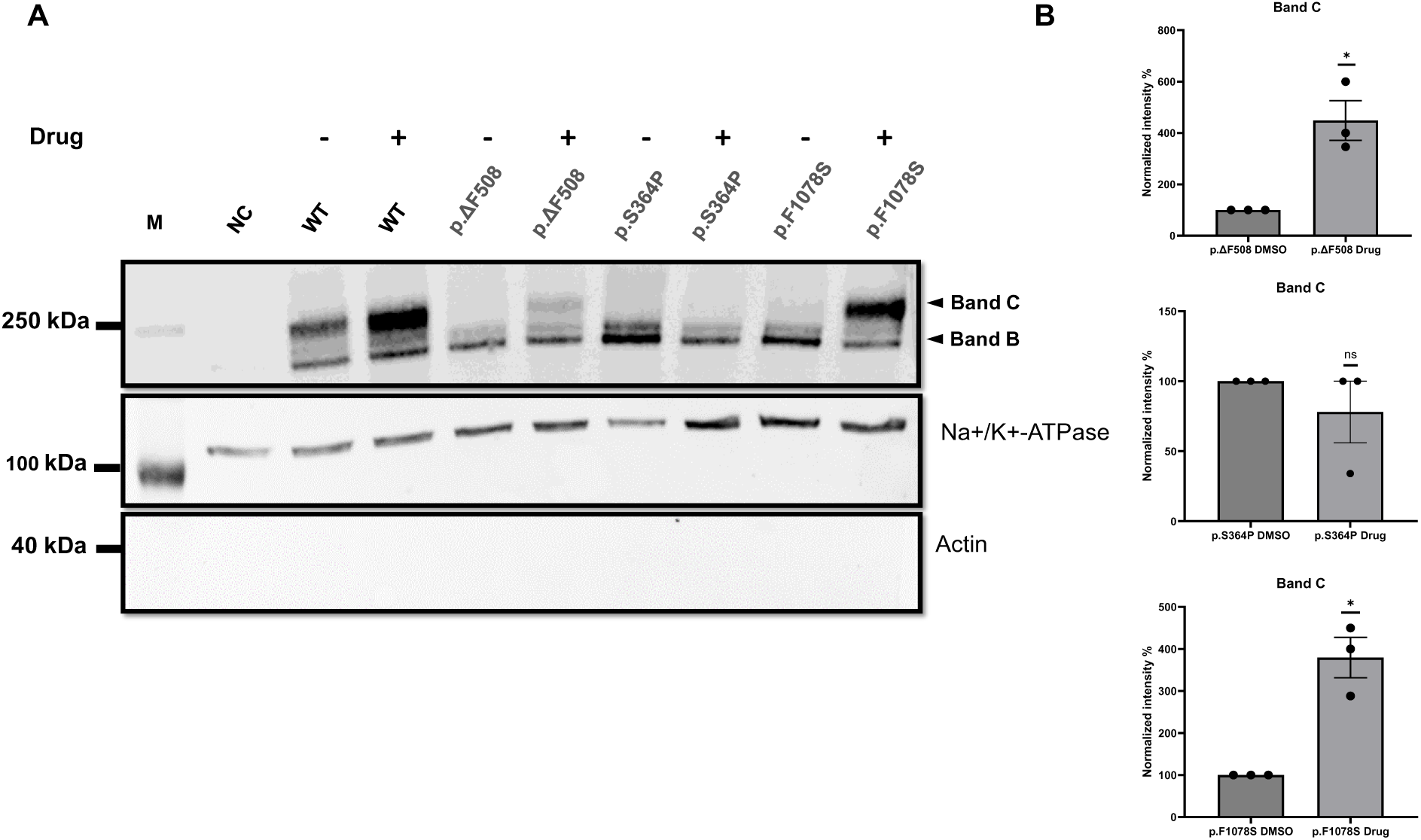
Immuno-blot of biotinylated experiments showing the expression of the wild-type CFTR (p.CFTR-WT) along with three variant forms: p.Phe508del (p.ΔF508), p.Ser364Pro(p.S364P), and p.Phe1078Ser(p.F1078S), treated (+) or not (-) was) with the Trikafta cocktail for 24 hours. **(B)** Quantification of three different blots performed as (A). Data are presented as mean ± SEM; \**p*<0.05; ns: non-significance, with p-value representing WT VS variant. M: Marker, NC: negative control.

## 4. Discussion

The principal findings of this study are the following: (1) it demonstrates the use of ONT for long-read sequencing of the complete *CFTR* gene by analysing samples from 9 individuals of Moroccan-African descent. This approach efficiently covered all coding and intronic regions, demonstrating the method’s effectiveness for detailed medical genetic studies. Specifically, using this method, all 6 pathogenic *CFTR* variants in the 3 probands with positive sweat tests could be identified. (2) The accuracy of ONT sequencing was validated against traditional Sanger sequencing, with all identified variants being confirmed. (3) The analytical pipeline we developed allowed phasing of the identified variants, which is important information in recessively inherited disorders like CF. (4) Biochemical characterisation revealed a significant reduction in the expression of the p.Ser364Pro and p.Phe1078Ser variants, indicating their potential role in CF pathology. Notably, the p.Phe1078Ser variant showed a positive response to the CFTR modulator drug treatment.

Our complete-gene sequencing approach is comparable to that of Salakhov *et al.* (for the genes *MYBPC3, MYH7, TPM1, TNNT2*, and *TNNI3*) and Soufi *et al.* (for the gene *LDLR*) [24], [25]. However, Soufi *et al.* did not aim to sequence the whole gene. Instead, they applied long-range PCR to amplify batches of exons, limiting subsequent identification of whole-gene haplotypes [25]. Complete-gene sequencing is essential, especially for genes with known disease-causing splice variants, as specific deep intronic variants can affect splicing [26]. Another advantage of the ONT-based method of this work is that it includes the possibility of multiplexing with manufacturer-provided standard barcodes (currently up to 96 indexes) [27]. In our study, we could multiplex the long-range PCR products of all individuals in one sequencing run. We did not try to multiplex more long-range PCR samples only because of the small number of samples.

The bioinformatics and data-processing tools available for ONT sequencing have significantly improved over the past few years. Several software tools are available for processing raw data, including Clair3 [28]. Usually, most bioinformatic protocols are included in EPI2ME Labs solutions from ONT or are recommended by them (https://labs.epi2me.io/). In our work, we developed a bioinformatic pipeline applying the Clair3 tool for calling SNVs and short indel variants. Using a combination of reference- and read-based phasing approaches, we could accurately phase the genotypes at these variants across the entire *CFTR* gene region. We also incorporated tools optimized for long-read sequencing data, Sniffles2 and cuteSV, to identify large structural variants accurately. However, in the small cohort of this study, we did not detect any significant structural variants. A recent study has suggested that LRS may improve the accuracy of diagnosing challenging, rare diseases by enabling the detection of structural variants, repeat expansions, and other unclear gene variants [29]. Therefore, for a rare disease such as CF with underdeveloped research, especially in Africa and low- and middle-income countries, implementing LRS to diagnose CF may improve its diagnostic rates. We propose that this new method can be an essential molecular screening tool for CF, especially in regions with limited access to genetic screening because of the significant genetic variation in Africa. It should be noted that while a cost analysis was not part of this study, it is very likely that the present ONT protocol is more cost-effective than the use of other NGS and long-read platforms. This affordability mainly results from the price of the sequencing device Mk1C (∼5,000 Euros) and the possibility of multiplexing the samples of many patients for whom the complete *CFTR* gene is sequenced.

Potential limitations of this study include the limited sample size, which compromises the methodology’s accuracy and applicability. To address this point, a more extensive CF-African cohort study will be initiated. The second limitation of this study is the lack of a control genotyped cohort from the Moroccan population, which will impact the assessment of the sensitivity and specificity of the current methodology.

## 5. Conclusion

In conclusion, this study provides evidence that ONT is an efficient and accurate method for identifying variants that cause CF. Our findings highlight the importance of sequencing the entire *CFTR* gene, which is especially crucial for populations with high genetic diversity, such as those found in African populations. Additionally, it underlines the need for biochemical and functional analyses to be conducted after identifying genetic variants. We propose that ONT-based sequencing could be the preferred method for diagnosing rare genetic diseases in countries with limited resources.

## Data Availability

The datasets used and/or analysed during the current study are available from the
corresponding author upon reasonable request.

## Abbreviations

CF: Cystic fibrosis
CFTR: CF transmembrane conductance regulator
HEK293: Human Embryonic Kidney 293
IGV: integrative genomics viewer
LRS: long-read sequencing
LR-PCR: Long-range polymerase chain reaction
NGS: next-generation sequencing
ONT: Oxford Nanopore Technology
SNP: single nucleotide polymorphism

## Acknowledgement

The authors thank the patients and their families for agreeing to participate in the study.

## Author contributions

**Nada El Makhzen:** Conceptualisation, Methodology, Investigation, Formal analysis, Validation, Writing - Original Draft, Writing - Review & Editing, Visualisation. **Alexander Nater:** Methodology, Validation, Writing - Review & Editing. **Alexandre Bokhobza:** Conceptualisation, Writing - Review & Editing. **Jean-Sébastien Rougier:** Conceptualisation, Writing - Review & Editing. **Anne-Flore Hämmerli:** Writing - Review & Editing. **Javier Sanz:** Formal analysis, Writing - Review & Editing. **Christiane Zweier:** Writing - Review & Editing. **Rémy Bruggmann:** Writing - Review & Editing. **Laila Bouguenouch:** Writing - Review & Editing. **Mounia Lakhdar Idrissi:** Writing - Review & Editing. **Hugues Abriel:** Supervision, Writing - Review & Editing.

## Availability of data and materials

The datasets used and/or analysed during the current study are available from the corresponding author upon reasonable request.

## Declarations

### Ethics approval and consent to participate

Approved by the Ethics Biomedical Research Committee, Mohammed V University, Rabat, Morocco.

### Consent for publication

--

